# Identification of circulating apolipoprotein M as a new determinant of insulin sensitivity and relationship with adiponectin

**DOI:** 10.1101/2023.02.09.23285709

**Authors:** Laurie Frances, Mikaël Croyal, Jean-Bernard Ruidavets, Marie Maraninchi, Guillaume Combes, Jérémy Raffin, Philippe de Souto Barreto, Jean Ferrières, Ellen E. Blaak, Bertrand Perret, Cédric Moro, René Valéro, Laurent O. Martinez, Nathalie Viguerie

## Abstract

**Background:** Adiponectin and apolipoprotein M (apoM) are adipokines indicatives of healthy adipose tissue and down-regulated with obesity. We compared circulating apoM with adiponectin regarding their relationship with metabolic parameters and insulin sensitivity and examined their gene expression patterns in adipocytes and in the adipose tissue.

**Methods:** Circulating apoM and adiponectin were examined in 169 men with overweight in a cross-sectional study, and 13 patients with obesity during a surgery-induced slimming program. Correlations with clinical parameters including the insulin resistance index (HOMA-IR) were analyzed. Multiple regression analyses were performed on HOMA-IR. The *APOM* and *ADIPOQ* gene expression were measured in the adipose tissue from 267 individuals with obesity and a human adipocyte cell line.

**Results:** Participants with type 2 diabetes had lower circulating adiponectin and apoM, while apoM was higher in individuals with dyslipidemia. Similar to adiponectin, apoM showed negative associations with HOMA-IR and hs-CRP (r>-0.2), and positive correlations with HDL markers (HDL-C and apoA-I, r>0.3). Unlike adiponectin, apoM was positively associated with LDL markers (LDL-C and apoB100, r< 0.20) and negatively correlated with insulin and age (r>-0.2). The apoM was the sole negative determinant of HOMA-IR in multiple regression models, while adiponectin not contributing significantly. After surgery, the change in HOMA-IR was negatively associated with the change in circulating apoM (r=-0.71), but not with the change in adiponectin. The *APOM* and *ADIPOQ* gene expression positively correlated in adipose tissue (r>0.44) as well as in adipocytes (r>0.81). In adipocytes, *APOM* was downregulated by inflammatory factors and upregulated by adiponectin.

**Conclusions:** The apoM rises as a new partner of adiponectin regarding insulin sensitivity. At the adipose tissue level, the adiponectin may be supported by apoM to promote a healthy adipose tissue.

## Introduction

Obesity is a global concern as excessive body fat is linked to metabolic syndrome, type 2 diabetes (T2D), and cardiovascular diseases [1]. Adipose tissue (AT) produces various factors, including numerous adipokines with local and broad systemic effects on whole-body homeostasis [2].

Most adipokines are secreted at higher levels with increasing fat and play a detrimental role in the development of metabolic disorders associated with obesity [2]. Conversely, few adipokines, like adiponectin, are inversely associated with body fat [2]. Adiponectin is an adipose-specific protein, encoded by *ADIPOQ*, which blood concentration decreases with obesity, metabolic syndrome, or T2D [3]. Adiponectin possesses anti-inflammatory, anti-diabetic, anti-atherogenic, and cardioprotective properties [4].

Recently, apolipoprotein M (apoM), predominantly produced by the liver and kidney, was found to be secreted by adipocytes [5]. Gene expression of *APOM* in AT is lower in individuals with high body fat, metabolic syndrome, or insulin resistance (IR) compared to lean healthy individuals [5]. Accordingly, blood levels of apoM are reduced in obesity, metabolic syndrome, and T2D [6-8]. Overall, apoM is an adipokine that exhibits a similar expression profile in AT and in the circulation to that of adiponectin [9]. Therefore, we hypothesized that both adiponectin and apoM could similarly predict changes in insulin sensitivity.

In this study, we conducted a comparative analysis of these two beneficial adipokines concerning obesity-related disorders, with a particular focus on glucose homeostasis. We investigated whether their blood concentrations were associated with lipid and glucose homeostasis parameters, and whether they could contribute to insulin sensitivity. Furthermore, we explored the relationship between apoM and adiponectin at the level of the adipocyte.

## Methods

### Study participants

The studies, referred as cohorts A, B and C, were conducted in accordance with the ethical principles of the Declaration of Helsinki. Before any procedure, all participants received oral information and signed a written informed consent form before they participated in any of the study procedures. Figure S1 illustrates the participants’ flowchart.

**Cohort A**: The protocol received approval from the local ethics committee (CCPPRB, Toulouse/Sud-Ouest, file #1-99-48, Feb 2000). The biological sample collection was registered as DC-2008-463 #1 to the Ministry of Research and to the Regional Health authority. Cohort A consisted of male participants selected from the Genetique et ENvironnement en Europe du Sud (GENES) cross-sectional case-control study. In this ancillary analysis, the patients with coronary artery disease (CAD) (referred to as cases) were age-matched to individuals without CAD (controls), and apoM was quantified using a multiplex liquid chromatography-tandem mass spectrometry (LC-MS/MS) method [10]. Subjects with missing data for fasting serum insulin and adiponectin levels were excluded, resulting in a final sample of 169 participants (mean age 60.6 years [SD, 8.1]), comprising 102 controls and 67 cases. Detailed procedures have been reported previously [11]. Anthropometric measurements, including fat mass assessment using Bioimpedance, and clinical variables were recorded. Venous blood was sampled after an overnight fast. Standardized interviews, conducted by a single physician, gathered information on environmental characteristics, medical history, and cardiometabolic risk factors. Alcohol consumption was calculated as the sum of different types of drinks. Physical activity was assessed using a standardized questionnaire and categorized into two levels: “high” if physical activity was performed for ≥ 20 min twice a week, or “low” if physical activity was performed for < 20 min once a week or less. Dyslipidemia was defined as treatment with drugs or fasting serum total cholesterol ≥ 2.50 g/L as described in [12]. Hypertension was defined as treatment with drugs or systolic blood pressure ≥ 140 mmHg or diastolic blood pressure ≥ 90 mmHg. T2D was defined as treatment with drugs or fasting blood glucose ≥ 7.0 mmol/L.

**Cohort B:** The approval was received by the Research Ethics Board of Aix-Marseille University (NCT01277068 and NCT02332434). Cohort B comprised 13 individuals with obesity of both sexes (77% women) with a mean age of 37.6 years [SD, 9.1], who underwent sleeve gastrectomy at the General and Endocrine Surgery Department of La Conception Hospital (Marseille, France), as previously described [13]. The clinical evaluation and laboratory tests were conducted at the Nutrition Department of La Conception Hospital. Patients were examined one month before (baseline) and one year after gastric sleeve surgery to reach steady weight loss. Each visit included measuring anthropometric parameters and blood sampling after an overnight fast [13].

**Cohort C:** This study is part of the Diet, Obesity, and Genes (DiOGenes) trial, which involved individuals with overweight or obesity recruited between 2005 and 2007 in 8 European countries [14]. The protocol received approval from the ethical boards of each of the 8 centers (NCT00390637). For the present study, a subgroup of 267 individuals (95 men and 172 women) with a mean age of 42.7 years [SD, 6.4]) was selected. Anthropometric measurements, blood sampling and subcutaneous abdominal AT needle biopsies were performed after an overnight fast. Clinical measures and subcutaneous abdominal AT samples were analysed at baseline (before any intervention), as described in [15].

### Biochemical analyses

Fasting blood glucose, triglycerides (TG), cholesterol, high-density lipoprotein-cholesterol (HDL-C), γ-glutamyltransferase (γ-GT), and high-sensitivity C-reactive protein (hs-CRP) were assayed with enzymatic reagents on an automated analyzer (Hitachi 912 and Cobas® Analyzer, Roche Diagnostics). Low-density lipoprotein-cholesterol (LDL-C) was calculated using the Friedewald formula. The estimated glomerular filtration rate (eGFR) was calculated using the Chronic Kidney Disease Epidemiology Collaboration (CKD-EPI) equation. Insulin concentration was measured by electrochemiluminescence immunoassay (Advia Centaur, Siemens Healthineers, cohort A and Cobas® Analyzer, Roche Diagnostics, cohort B). IR was assessed using the Homeostasis Model Assessment of Insulin Resistance (HOMA-IR) index, calculated as [fasting glucose (mM) x fasting insulin (mU/L)/22.5]. Adiponectin serum levels were measured using a specific enzyme-linked immunosorbent assay kit (Quantikine® Human Adiponectin, R&D Systems). Apolipoprotein A-I (apoA-I), apolipoprotein B100 (apoB100), and apolipoprotein C-III (apoC-III) were determined by immunoturbidimetric assay on an automated analyzer (Cobas® Analyzer, Roche Diagnostics, Cohort A) and by immunonephelometric assay (Roche Diagnostics) using a BN ProSpec analyzer (Siemens Healthineers). ApoM was measured by the LC-MS/MS method, as described previously [13].

### Cell culture

Cells were incubated at 37°C in a humidified atmosphere containing 5% CO2. Human multipotent adipose-derived stem cells (hMADS) were cultivated as described in [5]. Briefly, hMADS cells were differentiated for 13 days in 50/50 DMEM 1g/L glucose (Sigma-Aldrich) and Ham’s F-12 (Lonza) medium, with 125 nM transferrin, 10 nM insulin, 0.2 nM triiodothyronine, 100 μM 3-isobutyl-1-methylxanthine (IBMX), 1 μM dexamethasone (Sigma-Aldrich), and 100 nM rosiglitazone (Cayman). ThP-1 monocytes were cultured in RPMI-1640 medium containing 10% FBS, 1 mM sodium pyruvate, 50 µM 2-mercaptoethanol (Invitrogen), and 36 mM glucose (Sigma-Aldrich). For differentiation into M0 macrophages, the cells were seeded in a serum-free, 2-mercaptoethanol-free RPMI medium containing 100 nM 12-phorbol-13-myristate acetate (Sigma-Aldrich) for 48h. Macrophages were then polarized for 3 days, inducing the M2-like anti-inflammatory phenotype with 20 ng/mL IL-4 (Sigma-Aldrich) or the M1-like pro-inflammatory phenotype with 2 ng/mL Escherichia coli lipopolysaccharide and 10 ng/mL IFNγ (Sigma-Aldrich).

### Cell treatment

After 10 days of differentiation, the hMADS cells were treated with recombinant globular adiponectin (PeproTech), or CRP (Merck Millipore) or exposed to 50/50 conditioned media from M1-like or M2-like polarized ThP-1 for 72 hours. The cells were transduced with AAVs vectors (serotype 8) encoding either apoM or mCherry (designed and produced by the Vectorology platform of the Cancerology Research Center of Toulouse, Inserm UMR1037) for 72 hours, as described in [16]. Subsequently, cells were lysed using RLT buffer (Qiagen) containing 1% 2-mercaptoethanol (Sigma-Aldrich), flash frozen and stored at -80°C.

### Gene expression

RNA was extracted using a Rneasy mini kit (Qiagen) following manufacturer’s instructions. Total RNA was reverse transcribed using Superscript II reverse transcriptase (Invitrogen). Real-time PCR was performed using Taqman probes or SyberGreen primers assays with a QuantStudio 5 Real-Time PCR system (Applied Biosystems). For the AT samples, expression of *APOM* (Hs00219533_m1) or *ADIPOQ* (Hs00605917_m1) data were normalized to *PUM1* (Hs00472881_m1). For the hMADS cells, expression of *APOM* or *ADIPOQ* (forward GCAGAGATGGCACCCCTG, reverse GGTTTCACCGATGTCTCCCTTA) were normalized to *LRP10* (Hs01047362_m1).

### Statistical analyses

The categorical parameters are expressed as numbers (%) and the quantitative parameters as means ± the standard deviation (SD). Shapiro–Wilks test was used to examine the normality of distribution of residuals and Levene’s test to determine the homogeneity of the variances. When comparing cases to controls or baseline versus after surgery, the mean values of the quantitative variables were analyzed by Student’s *t*-test. When the basic assumptions of Student’s *t*-test were not satisfied, the data were logarithmically transformed or subjected to a Wilcoxon–Mann–Whitney test. Associations of apoM, adiponectin, and HOMA-IR with anthropometric parameters, biological markers, and environmental factors were tested using Spearman’s rank correlations with *P*-value adjustment using false discovery rate. For regression analyses of HOMA-IR, a backward stepwise selection was used. The F-statistics for a variable to stay in the model had to be significant at *P* < 0.10. Variables were maintained in the model when the F-statistic was significant at *P* < 0.05. For each quantitative variable, regression analyses were performed with polynomial models to look for possible nonlinear relationships with dependent variables. Lack of multicollinearity was assessed by examining the variance inflation factors in the final models.

## Results

### Circulating adiponectin versus apoM relationship with cardiometabolic status

The clinical and biological characteristics of cohort A are presented in Table S1. Cohort A comprised overweight men (BMI, 26.7 kg/m², [SD, 3.5]), of who 39.6% were patients with CAD (cases).

Compared with the control participants, the CAD patients had less physical activity and were diagnosed or treated more often for hypertension, dyslipidemia, or T2D, with a larger proportion suffering from T2D (25.4% versus 6.9% in the controls). The body fat mass percentage and waist circumference were also higher in cases than in the controls. Among the biological markers, the levels of total cholesterol and LDL-C were lower in cases, probably reflecting the effects of lipid-lowering drugs in patients. However, individuals with CAD displayed higher levels of TG, hs-CRP, and γ-GT. HDL markers (HDL-C and apoA-I) were lower. Finally, the cases exhibited higher HOMA-IR and lower adiponectin levels than the control participants, while the apoM levels were similar between the two groups.

Blood levels of apoM and adiponectin were then considered according to other disease conditions, including T2D, dyslipidemia, and hypertension (Table S2). Adiponectin and apoM levels were lower in participants diagnosed or treated for T2D. Unlike adiponectin, apoM levels were lower in participants with hypertension, while they were higher in subjects with dyslipidemia than in the individuals without these conditions. These observations were weakened when only the treatment conditions were considered (Table S3), except for adiponectin, for which the levels were lower in participants treated for dyslipidemia and T2D.

### Circulating apoM is a negative determinant of insulin resistance

Correlations between circulating levels of apoM and biological and clinical parameters relative to cardiovascular and T2D risk factors were investigated in the entire study population (cohort A). These correlations were compared with equivalent correlation for adiponectin and HOMA-IR (Table 1).

**Table 1.**
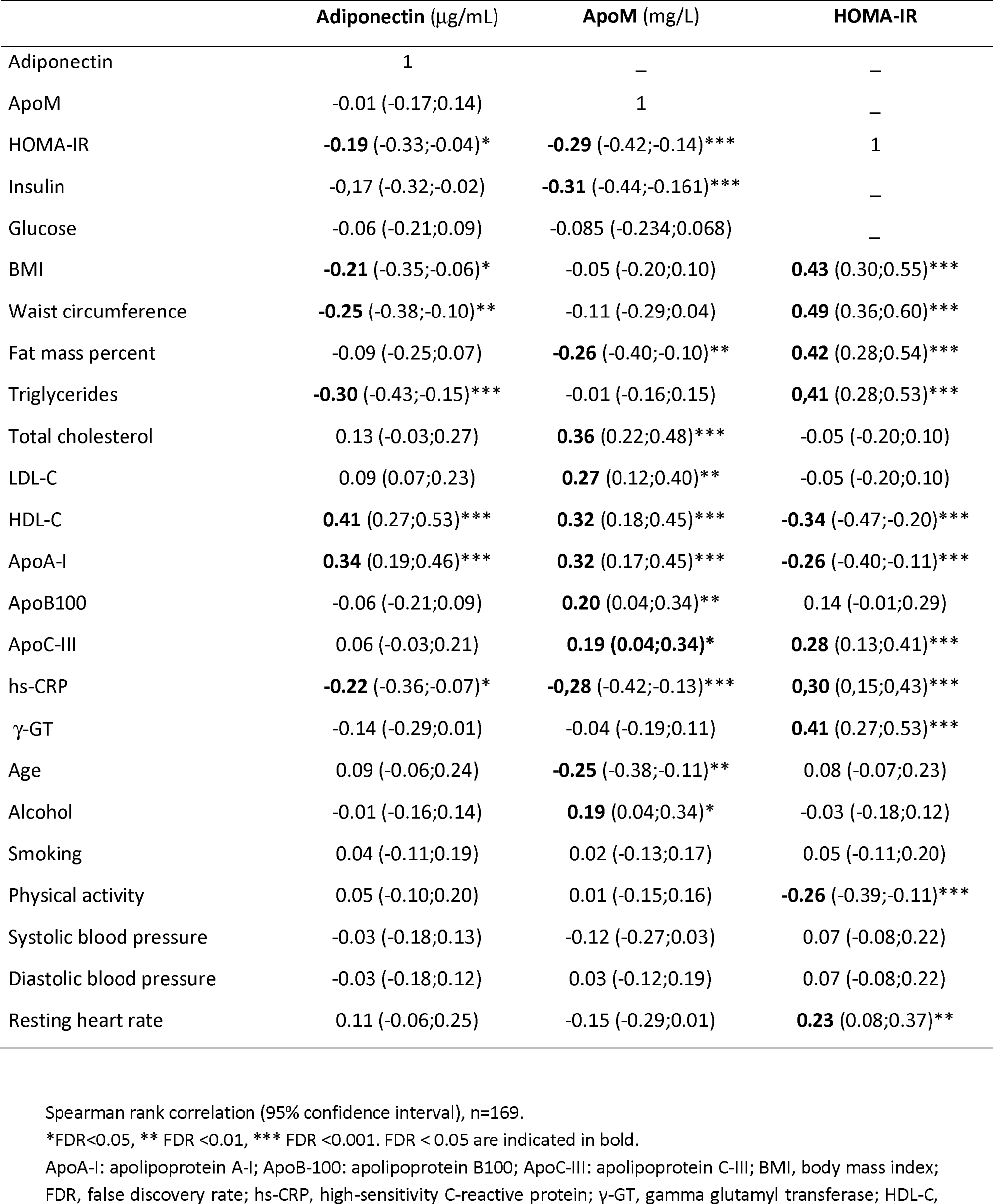
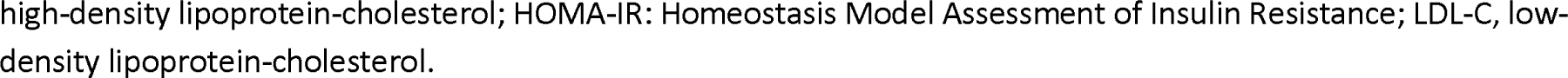
Spearman correlation coefficients of adiponectin, apoM and HOMA-IR with anthropometric parameters, biological markers and environmental factors in cohort A.

|  | Adiponectin (μg/mL) | ApoM (mg/L) | HOMA-IR |
| --- | --- | --- | --- |
| Adiponectin | 1 | — | — |
| ApoM | -0.01 (-0.17;0.14) | 1 | — |
| HOMA-IR | **-0.19** (-0.33;-0.04)* | **-0.29** (-0.42;-0.14)*** | 1 |
| Insulin | -0.17 (-0.32;-0.02) | **-0.31** (-0.44;-0.161)*** | — |
| Glucose | -0.06 (-0.21;0.09) | -0.085 (-0.234;0.068) | — |
| BMI | **-0.21** (-0.35;-0.06)* | -0.05 (-0.20;0.10) | **0.43** (0.30;0.55)*** |
| Waist circumference | **-0.25** (-0.38;-0.10)** | -0.11 (-0.29;0.04) | **0.49** (0.36;0.60)*** |
| Fat mass percent | -0.09 (-0.25;0.07) | **-0.26** (-0.40;-0.10)** | **0.42** (0.28;0.54)*** |
| Triglycerides | **-0.30** (-0.43;-0.15)*** | -0.01 (-0.16;0.15) | **0.41** (0.28;0.53)*** |
| Total cholesterol | 0.13 (-0.03;0.27) | **0.36** (0.22;0.48)*** | -0.05 (-0.20;0.10) |
| LDL-C | 0.09 (0.07;0.23) | **0.27** (0.12;0.40)** | -0.05 (-0.20;0.10) |
| HDL-C | **0.41** (0.27;0.53)*** | **0.32** (0.18;0.45)*** | **-0.34** (-0.47;-0.20)*** |
| ApoA-I | **0.34** (0.19;0.46)*** | **0.32** (0.17;0.45)*** | **-0.26** (-0.40;-0.11)*** |
| ApoB100 | -0.06 (-0.21;0.09) | **0.20** (0.04;0.34)** | 0.14 (-0.01;0.29) |
| ApoC-III | 0.06 (-0.03;0.21) | **0.19** (0.04;0.34)* | **0.28** (0.13;0.41)*** |
| hs-CRP | **-0.22** (-0.36;-0.07)* | **-0.28** (-0.42;-0.13)*** | **0.30** (0.15;0.43)*** |
| γ-GT | -0.14 (-0.29;0.01) | -0.04 (-0.19;0.11) | **0.41** (0.27;0.53)*** |
| Age | 0.09 (-0.06;0.24) | **-0.25** (-0.38;-0.11)** | 0.08 (-0.07;0.23) |
| Alcohol | -0.01 (-0.16;0.14) | **0.19** (0.04;0.34)* | -0.03 (-0.18;0.12) |
| Smoking | 0.04 (-0.11;0.19) | 0.02 (-0.13;0.17) | 0.05 (-0.11;0.20) |
| Physical activity | 0.05 (-0.10;0.20) | 0.01 (-0.15;0.16) | **-0.26** (-0.39;-0.11)*** |
| Systolic blood pressure | -0.03 (-0.18;0.13) | -0.12 (-0.27;0.03) | 0.07 (-0.08;0.22) |
| Diastolic blood pressure | -0.03 (-0.18;0.12) | 0.03 (-0.12;0.19) | 0.07 (-0.08;0.22) |
| Resting heart rate | 0.11 (-0.06;0.25) | -0.15 (-0.29;0.01) | **0.23** (0.08;0.37)** |

The blood level of apoM did not show any significant correlation with that of adiponectin. However, similar to adiponectin, apoM exhibited a negative association with hs-CRP (r=-0.28) and HOMA-IR (r=-0.29), while it was positively associated with HDL markers (r=0.32, for both HDL-C and apoA-I). Unlike adiponectin, apoM did not correlate with TG, but it was positively associated with total cholesterol (r=0.36), LDL markers (LDL-C and apoB100, r=0.27; and r=0.20, respectively), and apoC-III (r=0.19). No association was observed between apoM and cardiac functions (systolic blood pressure, heart rate), or environmental factors (smoking, physical activity), except for a positive correlation between apoM and alcohol consumption (r=0.19). Of note, apoM was negatively associated with fat mass (r=-0.26) and age (r=-0.28).

In addition to the expected negative correlations with apoM and adiponectin, HOMA-IR displayed negative correlations with HDL markers (apoA-I and HDL-C) and physical activity and positive correlations with adiposity markers (BMI, fat mass, and waist circumference), TG, apoC-III, hs-CRP, and γ-GT (Table1).

To further identify the determinants of IR among these variables associated with HOMA-IR, we conducted a multiple regression analysis with HOMA-IR as the dependent variable. The model included apoM and adiponectin as potent explanatory variables, along with BMI, waist circumference, physical activity, TG, HDL-C, hs-CRP, γ-GT, apoC-III, apoB100, T2D, dyslipidemia, hypertension, and case-control status. This model explained 39% of the variability in HOMA-IR and revealed that diabetes status and four variables were determinants of HOMA-IR variability, which included apoM but not adiponectin (Table 2). Among the identified determinants, waist circumference and γ-GT showed the strongest positive associations with HOMA-IR, explaining 13.6% and 9.6% of the variability, respectively. In contrast, apoM displayed an inverse association, accounting for 5.6% of the variability in HOMA-IR. Notably, no significant interaction between apoM and case-control status was observed (*P*=0.58). Furthermore, even after removing apoM from the set of variables at entry, adiponectin still did not contribute to the variability of HOMA-IR (data not shown).

**Table 2.** Multiple linear regression analysis on HOMA-IR in cohort A.

|  | R<sup>2</sup>=39 |  |  |  |
| --- | --- | --- | --- | --- |
| | $\beta$ estimate | SE | P-value | %<sup>b</sup> |
| ApoM | -0.133 | 0.045 | 0.0035 | 5.6 |
| Waist circumference<sup>a</sup> | 0.177 | 0.046 | 0.0002 | 13.6 |
| γ-GT<sup>a</sup> | 0.181 | 0.046 | 0.0001 | 9.6 |
| ApoC-III<sup>a</sup> | 1.421 | 0.044 | 0.0016 | 4.5 |
| Diabetes | 0.353 | 0.132 | 0.0084 | 2.8 |

Variables at entry: Adiponectin, apoC-III, apoB-100, apoM, BMI, γ-GT, hs-CRP, HDL-C, physical activity, triglycerides, waist circumference, diabetes (fasting blood glucose $\geq 7.0$ mmol/L or treatment vs. no), dyslipidemia (total cholesterol $\geq 2.50$ g/L or treatment), hypertension (systolic blood pressure $\geq 140$ mmHg or diastolic blood pressure $\geq 90$ mmHg or treatment) and case-control status. Continuous variables were standardised. $\beta$ estimates were given for one standard deviation variation.
<sup>a</sup>Log-transformed data; <sup>b</sup>Percentage of variance explained.
ApoB-100: apolipoprotein B100; ApoC-III: apolipoprotein C-III; ApoM: apolipoprotein M; BMI: body mass index; γ-GT: gamma-glutamyltransferase; HDL-C: high-density lipoprotein cholesterol; hs-CRP: high-sensitivity C-Reactive Protein; HOMA-IR, Homeostasis Model Assessment of Insulin Resistance; LDL-C: low-density lipoprotein cholesterol.

### Increased circulating apoM is closely related to insulin resistance relief after sleeve gastrectomy

To further study the association between apoM and IR, we examined the changes in circulating apoM after sleeve gastrectomy in patients with obesity and analyzed their correlation with the improvement in HOMA-IR. Anthropometric and biochemical characteristics of these patients at baseline (*i.e.,* before gastric sleeve surgery) and one year after surgery are presented in Table S4. One year after surgery, all individuals showed a decrease in adiposity markers (BMI, fat mass, and waist circumference) and displayed an improved lipid profile, including reduced atherogenic lipids (TG and LDL-C) and increased HDL markers (HDL-C and apoA-I). Additionally, there was a significant decrease in HOMA-IR (from 4.26, [SD, 1.49] to 1.39, [SD, 0.69]) and an increase in adiponectin levels (from 5.54 µg/L, [SD, 1.61] to 10.47 µg/L [SD, 4.07]). Circulating levels of apoM were not significantly changed (Table S4); however, we found a strong inverse relationship between the changes in HOMA-IR and changes in circulating apoM (r=-0.71), while no significant association was observed with changes in adiponectin (Figure 2). Of note, there was a trend of a positive correlation between changes in circulating apoM and changes in adiponectin (r=0.57, *P*=0.08, not shown).

**Figure 1.**
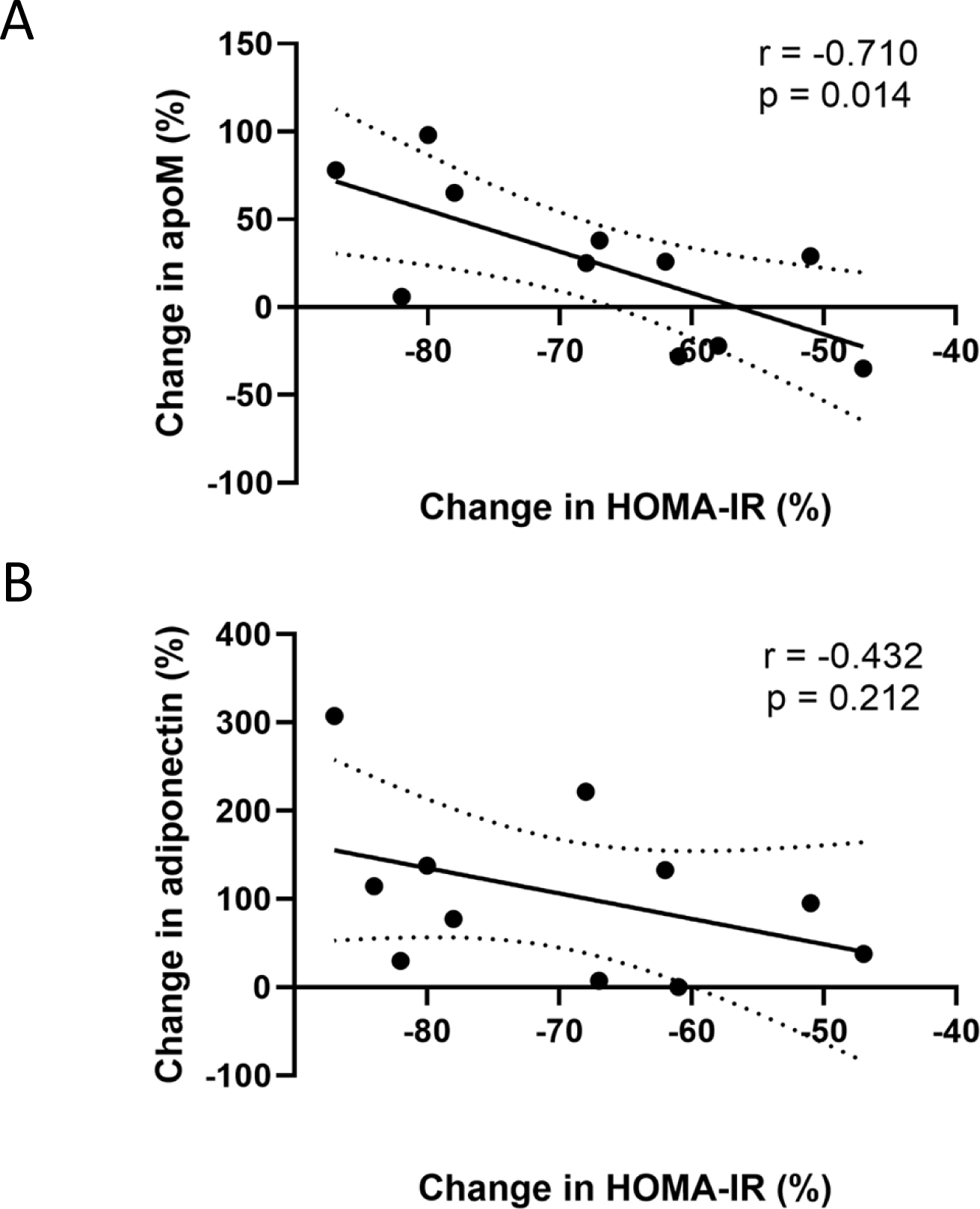
Changes in circulating apoM and adiponectin relationship with improvement of insulin sensitivity after bariatric surgery. Sera from 11 obese individuals (cohort B) were tested for apoM (panel A) and adiponectin (panel B) levels before and one year after sleeve gastrectomy. Changes in adipokine concentrations were correlated to the changes in the Homeostasis Model Assessment of Insulin Resistance (HOMA-IR) index. Data are expressed as percentage change from baseline. Linear correlation coefficients and *P-*values are displayed in each graph. The dotted lines represent 95% confidence intervals.

**Figure 2.**
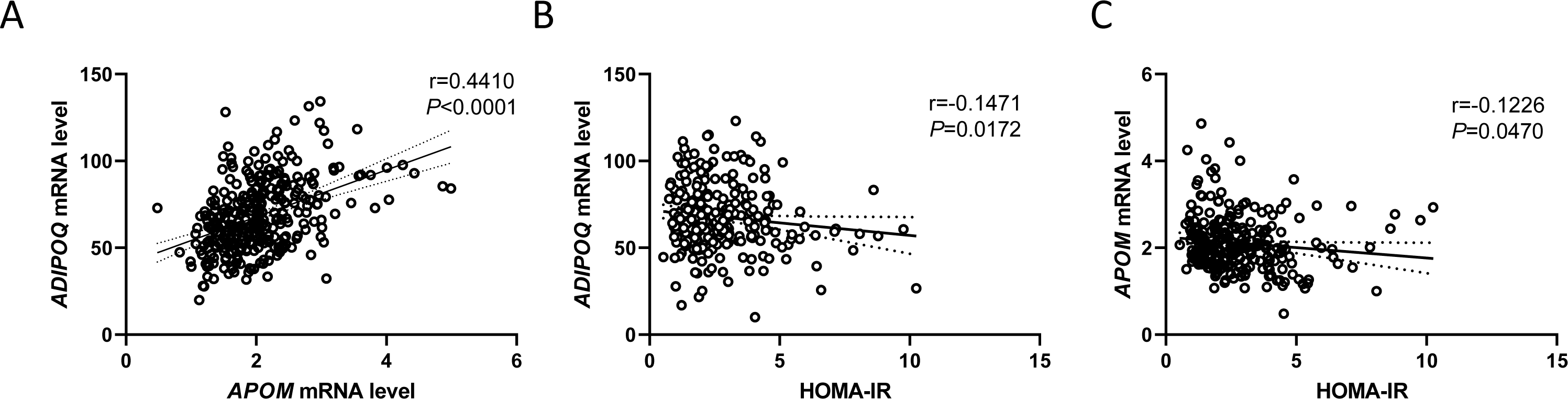
*ADIPOQ* and *APOM* gene expression in adipose tissue from overweight and obese individuals. *ADIPOQ* and *APOM* mRNA levels were measured in subcutaneous adipose tissue of 267 men and women with overweight or obesity (cohort C). Linear regression analysis of *ADIPOQ* and *APOM* gene expression (panel A). Linear regression of *APOM* (panel B) and *ADIPOQ* (panel C) gene expression to HOMA-IR. Linear correlation coefficient and *P*-value are presented. Dotted lines represent 95% confidence interval.

### Positive association between ADIPOQ and APOM mRNA level in AT

We compared the gene expression of adiponectin and apoM in human subcutaneous AT from individuals with overweight or obesity (cohort C, Figure 2A and Table S5). *ADIPOQ* gene expression in AT was higher than *APOM* gene expression and both were positively correlated (Figure 2A, r=0.441). Additionally, HOMA-IR negatively correlated with AT expression of *ADIPOQ* (r=0.147, Figure 2B), and *APOM* gene expression in AT (r=-0.1226, Figure 2C).

### Inflammatory factors downregulate ADIPOQ and APOM expression in adipocytes

Due to the close association of IR with AT inflammation, we aimed to mimic the impact of inflammatory conditions on *ADIPOQ* and *APOM* expression in adipocytes. We treated hMADS adipocytes with conditioned media from pro-inflammatory M1-like or anti-inflammatory M2-like polarized ThP-1 macrophages. The former media led to down-regulation of both *ADIPOQ* and *APOM* gene expressions (Figure 3A). A strong correlation between *ADIPOQ* and *APOM* gene expression was observed in these adipocytes (r=0.91, Figure 3B). Similarly, a treatment with CRP also downregulated the expression of *ADIPOQ* and *APOM* genes in hMADS cells (Figure 3C), while the positive correlation between *ADIPOQ* and *APOM* mRNA levels persisted under this condition (r=0.82, Figure 3D).

**Figure 3.**
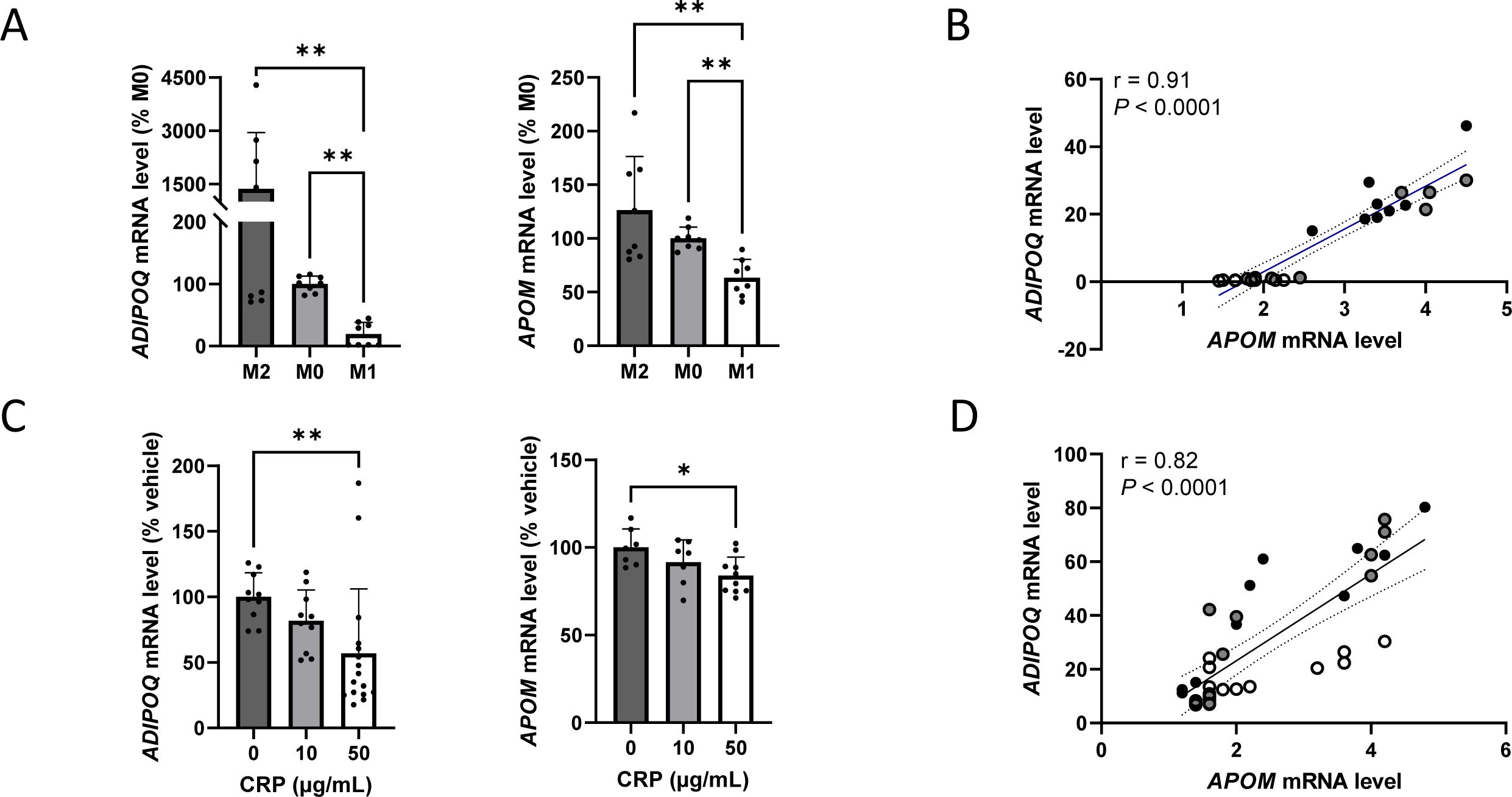
Inflammatory factors downregulate *ADIPOQ* and *APOM* gene expression in adipocytes. Effect of conditioned media from non-polarized, M2-like polarized or M1-like polarized ThP-1 cells on *ADIPOQ* gene expression in hMADS adipocytes (panel A). Association between *ADIPOQ* and *APOM* gene expression in hMADS adipocytes treated for 48 h with conditioned media from non-polarized (grey dots), M2-like polarized (black dots) or M1-like polarized (white dots) ThP-1 cells (panel B). *ADIPOQ* and *APOM* gene expression in hMADS adipocytes after C-reactive protein treatment for 48 h (panel C). Association between *ADIPOQ* and *APOM* gene expression in hMADS adipocytes treated for 48 h with C-reactive protein (panel D). Results are presented as mean ± SD of 3 independent experiments (panels A and C). Linear correlation coefficients and *P-*values are displayed in each graph. The dotted lines represent 95% confidence intervals (panels B and D). Data were analysed by Kruskal-Wallis test (panels A and C). \**P*<0.05. \*\**P*<0.01.

### Adiponectin promotes APOM expression in adipocytes

Adipocytes express specific adiponectin receptors, whereas apoM lacks known specific receptor. To further investigate the relationship between the two adipokines, we examined the effect of adiponectin on *APOM* gene expression. Treating hMADS adipocytes with increasing concentrations of adiponectin resulted in a dose dependent increase in *APOM* expression, with a mean 1.6-fold [SD, 0.9]) rise at 1µM adiponectin (Figure 4).

**Figure 4.**
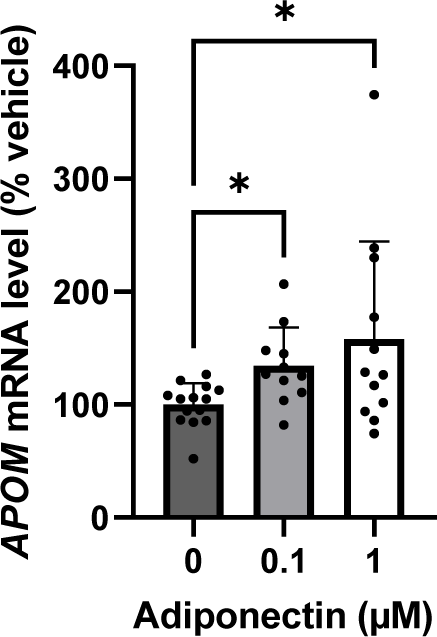
Adiponectin upregulates *APOM* gene expression in adipocytes. *APOM* mRNA level upon 48 h of globular adiponectin treatment in hMADS adipocytes. Results are presented as mean ± SD of 3 independent experiments. Data were analysed by Kruskal-Wallis test. \**P*<0.05. \*\**P*<0.01.

## Discussion

Here, we explored the relationship between the circulating concentrations of two beneficial adipokines, namely adiponectin and apoM, and cardiometabolic parameters in high-risk populations. We also examined the relationship between the two adipokines at the adipose level.

Firstly, despite the strong correlation of *APOM* and *ADIPOQ* gene expression in adipocytes and AT, the apoM concentration did not correlate with that of adiponectin, as a hallmark of their distinct tissue origin. While their respective circulating levels were lower in participants with T2D, their level may still reflect distinct cardiometabolic states.

As anticipated, our study validated previously reported associations between blood levels of adiponectin and lipids [17, 18]. Adiponectin displayed an inverse correlation with TG and a positive association with HDL markers such as HDL-C and apoA-I. In contrast, the circulating apoM level was not associated with plasma TG but displayed a positive correlation with both anti-atherogenic HDL (HDL-C, apoA-I) and atherogenic LDL (LDL-C, apoB100) markers. This association of apoM with both HDL and LDL markers has been reported previously [6, 19] and is likely due to the fact that apoM is transported by both HDL and LDL rather than having a specific role in lipoprotein metabolism [19]. However, it is important to note that apoM is predominantly transported by HDL, and a recent report has shown that low plasma apoM is independently associated with heart failure [20]. This suggest that apoM may play a role in some HDL cardioprotective functions [9].

An expected reduced level of adiponectin was found in patients with CAD [17]. In humans, the *ADIPOQ* gene, located on chromosome 3q27, contains polymorphisms that influence the level and activity of adiponectin, SNP 276 (G>T) and I164T. The former is associated with obesity, IR, and T2D [21], while the latter is a missense mutation associated with metabolic syndrome and CAD [22]. There is consistent evidence of an inverse relationship between plasma adiponectin and metabolic syndrome, low-grade inflammation, T2D, and cardiovascular risk [23-25]. Of note, circulating adiponectin has paradoxically been associated with increased all-cause and cardiovascular mortality [26, 27]. Adiponectin resistance in vessels might be a clue to explain this unexpected outcome on the prognostic value of circulating adiponectin [27].

ApoM concentration was found to be higher in participants with dyslipidemia, consistent with previous studies [8]. This observation aligns with the previously mentioned positive association of apoM with LDL-C. Interestingly, the relationship between apoM and dyslipidemia status was no longer observed when only the treated patients (the majority of whom were on statin treatment) were considered among the subjects with dyslipidemia. This is consistent with the effect of statins in lowering apoM levels [6]. Additionally, apoM levels were lower in subjects with hypertension, in accordance with its role in reducing blood pressure in hypertensive mice [28].

Similarly to *APOM* expression in AT [5] and consistent with previous reports [18, 20], circulating apoM was found to be negatively associated with body fat and hs-CRP level, a biomarker of systemic inflammation. Additionally, our *in vitro* studies on adipocytes indicate that conditions mimicking an inflammatory context, including treatments with conditioned media from pro-inflammatory macrophages or CRP, down-regulate *APOM* gene expression. This is in line with our previous study reporting a negative effect of TNFα on *APOM* gene expression in adipocytes [5]. This suggests that a low circulating apoM may serve as a potential marker of systemic inflammation and dysfunctional AT.

While adiponectin is a key adipokine in preventing metabolic syndrome and associated pathologies, much less is known about apoM, especially regarding its expression in adipocytes. ApoM is distributed in various tissues, with primary expression in hepatocytes and proximal tubular kidney cells, and to a lesser extent in endothelial cells and adipocytes [9, 29]. The human *APOM* gene is located on chromosome 6p21.33. There is evidence that T-778C polymorphism in the proximal promoter region of the *APOM* gene confers susceptibility to the development of T2D [30]. Another variant, rs1266078, resulted in an 11% lower circulating apoM, but no association was found with the risk of T2D [31]. Similar to adiponectin, we found a lower level of apoM in participants with T2D and an inverse association with IR. Surprisingly, in a multiple linear regression model, the apoM level in cohort A was the only negative determinant of HOMA-IR, independently of coronary status, while adiponectin did not contribute to the variability of HOMA-IR. This indicates that in men with overweight, blood apoM has a stronger association to HOMA-IR than circulating adiponectin.

To investigate the relationship between these two adipokines and IR, we examined whether weight-reduction surgery in patients with obesity (cohort B) could influence the levels of apoM and its association with IR. As reported elsewhere, patients with obesity (cohort B, baseline) exhibited lower circulating levels of adiponectin [32] and apoM [33] compared to individuals with overweight (cohort A), consistent with the previously mentioned negative association of plasma adiponectin and apoM with fat mass. Bariatric surgery offers an efficient approach for managing obesity, with various surgical procedures available [34]. Among these, sleeve gastrectomy accounts for 55.4% of all bariatric surgeries thanks to its minimal invasiveness [35] and has been shown to resolve IR and T2D [36]. In our study, one year after sleeve gastrectomy, all patients exhibited a HOMA-IR below 2.5 and a two-fold increase in adiponectin concentration. The minor increase (+13%) in apoM levels did not reach statistical significance (*P* > 0.1). Circulating apoM has been reported to be filtered then taken up from urine by tubular kidney cells *via* the megalin receptor [37]. Given that people with class 3 obesity (BMI>40 kg/m^2^) often present kidney disease that could be alleviated after sleeve gastrectomy, one might expect an increase in plasma apoM levels associated with improved renal function. However, the patients of Cohort B had mild CKD as measured by eGFR, and no significant change in eGFR was found, in line with a recent report [38]. Conversely, improvement in insulin sensitivity following surgery was strongly associated with increase in circulating apoM. This reciprocal correlation between apoM and HOMA-IR suggests that increased circulating apoM could improve glucose homeostasis by reducing IR. This provides additional evidence that, in complement to adiponectin, apoM may be an additional beneficial factor of glycaemic control.

Next, we examined the relationship between adiponectin and apoM at the AT level. For the first time, we reported here a strong positive correlation between *ADIPOQ* and *APOM* gene expression in human AT and adipocytes, independently of the inflammatory context. Interestingly, adiponectin was found to promote the expression of *APOM* in adipocytes. Although further studies are needed to identify the pathways regulating apoM expression and secretion in adipocytes, our observations suggest a positive feedback loop between adiponectin and apoM that controls local AT inflammation and might help prevent insulin sensitivity decline. Considering the importance of AT inflammation in systemic IR [4], our study suggests that the adipose apoM might play a noticeable role in maintaining a healthy AT environment and promoting systemic insulin sensitivity, alongside with adiponectin.

Several limitations of this study warrant mention. Firstly, we focused solely on one form of circulating adiponectin. Considering that the high molecular weight isoform is considered as the active form [32], further investigations to explore isoform specificity would be necessary. Secondly, additional studies are required to determine if AT is a significant site of apoM production and to understand the mechanism through which apoM may contribute to preventing IR.

In conclusion, the present study examined for the first time the relative contributions of adiponectin and apoM regarding insulin sensitivity. We found a close relationship between adipose apoM and adiponectin, and that plasma apoM, but not adiponectin, was a negative explanatory variable of HOMA-IR, independently of coronary status. The stronger relationship of insulin sensitivity with apoM rather than adiponectin was unexpected. This establishes apoM as a novel factor associated to insulin sensitivity.

## Acknowledgments

We are grateful to all individuals participating in this study. This research was funded by the Region Occitanie Pyrénées-Méditerranée (GRAINE HEPATOCARE, Reference number: no. 19014226), Inserm, Toulouse 3 University, and grants from Société Française de Nutrition. L.F. was supported by a Ph.D. fellowship from Region Occitanie Pyrénées-Méditerranée and Inserm.

## Author Contributions

Conceptualization, L.O.M. and N.V.; methodology, LO.M., L.F., G.C., J.F., B.P., E.E.B, R.V., M.C., and N.V.; formal analysis, J.B.R. and M.M.; data curation, L.F., J.B.R., J.R., and M.C.; draft preparation, L.O.M. and N.V.; review and editing, L.F., J.B.R., M.M., G.C., J.R, P.S.R., J.F.; B.P.; C.M.; R.V.; M.C., E.E.B, L.O.M, and N.V.; supervision, L.O.M. and N.V.; funding acquisition, L.O.M. and N.V..

## Data Availability

The datasets analyzed during the current study are not publicly available but are available from the corresponding authors on reasonable request.

https://figshare.com/s/04d2de1ad0fd8385fc88

