## Supplementary Information for "Identification of circulating apolipoprotein M as a new determinant of insulin sensitivity and relationship with adiponectin"

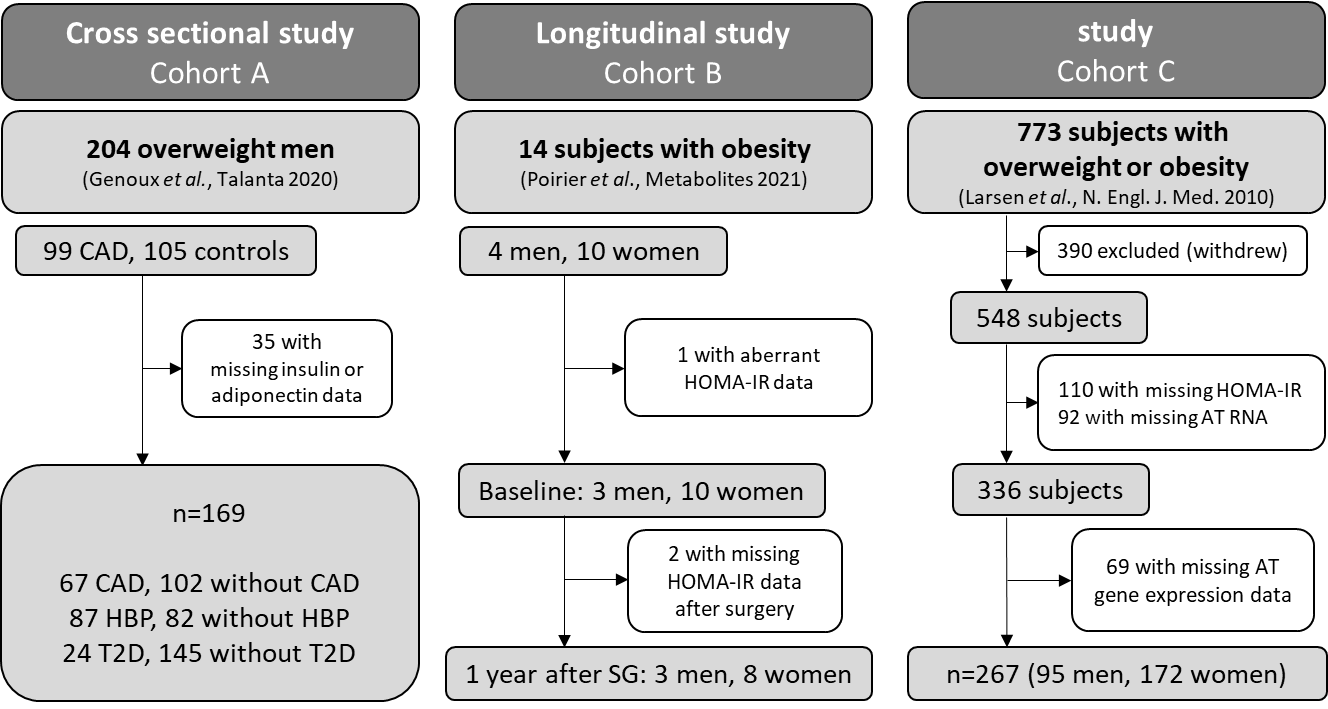

**Figure S1. Flowchart of participants.** AT: adipose tissue; CAD: coronary artery disease; HBP: High blood pressure (systolic blood pressure ≥140 mmHg or diastolic blood pressure ≥90 mmHg or treatment); HOMA-IR: Homeostasis Model Assessment of Insulin Resistance; SG: sleeve gastrectomy; T2D: type 2 diabetes (fasting blood glucose ≥7.0 mmol/L or treatment).

| **Table S1. Clinical and biological characteristics in cohort A, comparing subjects with and without coronary artery disease** | | | | | | | | | | | |
| --- | --- | --- | --- | --- | --- | --- | --- | --- | --- | --- | --- |
|  | | | **Whole population**  n = 169 (100%) | | **Controls (No CAD)**  n = 102 (60.4%) | | | **Cases (CAD)**  n = 67 (39.6%) | | ***P-*value** | |
| Age, year | | 60.6 (8.1) | | | 60.9 (8.2) | | | 60.2 (7.9) | | 0.59 | |
| BMI, kg/m^2^ | | 26.7 (3.5) | | | 26.4 (3.2) | | | 27.1 (3.8) | | 0.19 | |
| **Fat mass, % body weight** | | 27.1 (5.3) | | | 26.4 (5.1) | | | 28.2 (5.5) | | **0.037** | |
| **Waist circumference, cm** | | 96.3 (9.8) | | | 94.4 (9.2) | | | 99.1 (10.1) | | **0.02** | |
| **Triglycerides, g/L^b^** | | 1.29 (0.72) | | | 1.09 (0.55) | | | 1.61 (0.83) | | **0.001** | |
| **Total cholesterol, g/L** | | 2.14 (0.39) | | | 2.24 (0.36) | | | 1.98 (0.4) | | **0.001** | |
| **LDL-C, g/L** | | 1.36 (0.35) | | | 1.44 (0.33) | | | 1.24 (0.36) | | **0.001** | |
| **HDL-C, g/L** | | 0.52 (0.15) | | | 0.58 (0.13) | | | 0.42 (0.12) | | **0.001** | |
| **ApoA-I, g/L** | | 1.42 (0.29) | | | 1.55 (0.25) | | | 1.22 (0.23) | | **0.001** | |
| ApoB100, g/L | | 1.05 (0.23) | | | 1.05 (0.21) | | | 1.04 (0.25) | | 0.21 | |
| ApoC-III, mg/L | | 30.7 (11.2) | | | 30.0 (10.3) | | | 31.9 (12.3) | | 0.27 | |
| Fasting glucose, mmol/L | | 5.6 (1.5) | | | 5.4 (1.3) | | | 5.8 (1.8) | | 0.69^a^ | |
| **Insulin, IU/L** | | 11.3 (11.4) | | | 9.8 (6.5) | | | 13.7 (16) | | **0.02^a^** | |
| **HOMA-IR** | | 2.9 (3.3) | | | 2.51 (2.7) | | | 3.53 (3.96) | | **0.008** | |
| **Adiponectin, μg/mL** | | 7.2 (5.1) | | | 8.4 (5.4) | | | 5.4 (4.1) | | **0.001** | |
| ApoM, mg/L | | 25.8 (7.2) | | | 25.9 (6.4) | | | 25.5 (8.3) | | 0.42 | |
| **hs-CRP, mg/L^b^** | | 8.4 (16.5) | | | 2.9 (5.4) | | | 17 (23) | | **0.001** | |
| **γ-GT, IU/L** | | 51.5 (58.4) | | | 43 (44.7) | | | 64.4 (73.1) | | **0.01^a^** | |
| Resting heart rate, bpm | | 62.7 (10.7) | | | 62.3 (9.5) | | | 63.4 (12.3) | | 0.52 | |
| Systolic blood pressure, mm Hg | | 137 (17) | | | 137 (16) | | | 138 (18) | | 0.82 | |
| Diastolic blood pressure, mmHg | | 83 (8) | | | 82 (7) | | | 83 (10) | | 0.44 | |
| Alcohol, g/day | | 26.1 (24.2) | | | 25.7 (21.4) | | | 26.7 (28.1) | | 0.49^a^ | |
| **Smoking, cigarettes/day** | | 2.1 (6.2) | | | 1.6 (6.1) | | | 2.9 (6.3) | | **0.027^a^** | |
| **Physical activity, high level, %^c^** | | 29.6 | | | 44.1 | | | 7.5 | | **0.001** | |
| **Diabetes, %^d^** | | 14.2 | | | 6.9 | | | 25.4 | | **0.001** | |
| **Treatment diabetes, %** | | 11.8 | | | 4.9 | | | 22.4 | | **0.001** | |
| **Dyslipidemia, %^e^** | | 55 | | | 47.1 | | | 67.2 | | **0.01** | |
| **Treatment dyslipidemia, %** | | 40.8 | | | 24.5 | | | 65.7 | | **0.001** | |
| **Hypertension, %^f^** | | 51.5 | | | 43.1 | | | 64.2 | | **0.007** | |
| **Treatment hypertension, %** | | 31.9 | | | 22.6 | | | 46.3 | | **0.001** | |
| Variables were measured at baseline (i.e. when individuals were first included in the GENES cohort). For continuous variables, values are expressed as means (SD). For categorical variables, values are expressed as number with frequency (%) in parentheses. Paired Student’s t-test, unless otherwise mentioned. *P*-values < 0.05 are indicated in bold.  ^a^Kruskal-Wallis test; ^b^Log-transformed data; ^c^“High” physical activity for 20 min at least twice a week versus “low” physical activity once a week or less; ^d^Diabetes, fasting blood glucose ≥7.0 mmol/L or treatment; ^e^Dyslipidemia, total cholesterol ≥2.50 g/L or treatment; ^f^Hypertension, systolic blood pressure ≥140 mmHg or diastolic blood pressure ≥90 mmHg or treatment; BMI: body mass index; Bpm: beats per minute; CAD: coronary artery disease; HDL-C: high-density lipoprotein cholesterol; hs-CRP: high-sensitivity C-reactive protein; γ-GT: Gamma-glutamyltranspeptidase; HOMA-IR: Homeostasis Model Assessment of Insulin Resistance; LDL-C: low-density lipoprotein cholesterol. | | | | | | | | | | | |
| **Table S2. ApoM and Adiponectin levels in cohort A according to cardiometabolic status** | | | | | | | | | | | |
|  |  | **n** | | **ApoM**  (mg / L) | | ***P*-value** |  | | **Adiponectin**  (μg / L)^a^ | | ***P*-value** |
|  | **Coronary artery disease** |  | |  | |  |  | |  | |  |
|  | no | 102 | | 25.9 (6.4) | | 0.42 |  | | 8.4 (5.4) | | **0.001** |
|  | yes | 64 | | 25.5 (8.3) | |  |  | | 5.4 (4.1) | |  |
|  | **Hypertension**^b^ |  | |  | |  |  | |  | |  |
|  | no | 82 | | 27.1 (6.7) | | **0.021** |  | | 7.7 (4.6) | | 0.26 |
|  | yes | 87 | | 24.5 (7.3) | |  |  | | 6.8 (5.6) | |  |
|  | **Dyslipidemia**^c^ | | | | | | | | | | |
|  | no | 76 | | 23.9 (6.2) | | **0.0017** |  | | 7.8 (4.9) | | 0.21 |
|  | yes | 93 | | 27.3 (7.5) | |  |  | | 6.7 (5.4) | |  |
|  | **Type 2 diabetes**^d^ | | | | | | | | | | |
|  | no | 145 | | 26.3 (7.1) | | **0.026** |  | | 7.6 (5.3) | | **0.008** |
|  | yes | 24 | | 22.8 (7.1) | |  |  | | 4.9 (3.4) | |  |
|  | ApoM and adiponectin were measured at baseline (i.e. when individuals were first included in the GENES cohort) and expressed as mean (SD).  Paired Student’s t-test; P-values < 0.05 are indicated in bold.  ^a^Log-transformed data.  ^b^Hypertension, systolic blood pressure ≥140 mmHg or diastolic blood pressure ≥90 mmHg or treatment;  ^c^Dyslipidemia, total cholesterol ≥2.50 g/L or treatment.  ^d^Diabetes, fasting blood glucose ≥7.0 mmol/L or treatment. | | | | | | | | | | |

| **Table S3. ApoM and adiponectin according to treatments in the study population (n = 169)** | | | | | | |
| --- | --- | --- | --- | --- | --- | --- |
|  | n | **ApoM**  (mg / L) | p for  treatment effect |  | **Adiponectin**  (μg / mL)^a^ | p for  treatment effect |
| **Treatment for hypertension** no | 115 | 25.8 (6.9) | 0.91 |  | 7.7 (5.1) | 0.49 |
| yes | 54 | 25.7 (7.7) |  |  | 6.1 (5.1) |  |
| *Beta-blocker agents* no | 137 | 25.8 (7.1) | 0.72 |  | 7.4 (5.2) | 0.22 |
| yes | 32 | 25.3 (7.3) |  |  | 6.2 (4.9) |  |
| *ACE inhibitor* no | 150 | 26.0 (7.1) | 0.18 |  | 7.3 (5.0) | 0.61 |
| yes | 19 | 23.7 (7.3) |  |  | 6.6 (6.3) |  |
| *Calcium channel inhibitors* no | 157 | 25.8 (7.1) | 0.59 |  | 7.2 (5.2) | 0.76 |
| yes | 12 | 24.7 (7.8) |  |  | 7.7 (5.0) |  |
| **Treatment for diabetes** no | 149 | 26.1 (7.1) | 0.14 |  | 7.5 (5.3) | 0.023^a^ |
| yes | 20 | 23.5 (7.3) |  |  | 4.7 (2.1) |  |
| *Insulin* no | 158 | 25.8 (7.0) | 0.65 |  | 7.4 (5.3) | 0.27 ^a^ |
| yes | 11 | 24.8 (9.3) |  |  | 5.1 (2.1) |  |
| *Metformin* no | 165 | 25.8 (7.3) | 0.58 |  | 7.3 (5.2) | 0.26 |
| yes | 4 | 23.5 (2.9) |  |  | 4.3 (2.4) |  |
| *Sulphonylureas* no | 162 | 25.8 (7.2) | 0.58 |  | 7.3 (5.2) | 0.14 ^a^ |
| yes | 7 | 24.3 (6.2) |  |  | 4.3 (2.0) |  |
| **Treatment for dyslipemia** no | 100 | 24.9 (6.4) | 0.08 |  | 7.9 (5.0) | 0.026 |
| yes | 69 | 27.0 (8.0) |  |  | 6.1 (5.3) |  |
| *Statin* no | 109 | 24.2 (6.6) | 0.35 |  | 8.1 (5.3) | 0.49 |
| yes | 60 | 25.5 (7.9) |  |  | 7.3 (6.5) |  |
| *Fibrate* no | 157 | 25.7 (7.1) | 0.93 |  | 7.3 (5.2) | 0.31 |
| yes | 12 | 25.9 (8.6) |  |  | 5.8 (4.1) |  |

Data are expressed as means (SD).

Paired Student’s t-test, unless otherwise mentioned.

^a^Log-transformed data.

| **Table S4. Anthropometric, biological and clinical characteristics at baseline and one year after sleeve gastrectomy in cohort B.** | | | | | |
| --- | --- | --- | --- | --- | --- |
|  | **Baseline** | **n** | **One-year after sleeve gastrectomy** | **n** | ***P*-value** |
| **BMI**, kg/m^2^ | 42.2 (4.6) | 13 | 28.5 (3.9) | 13 | **<0.001** |
| **Fat mass**, % body weight | 49.8 (5.3) | 13 | 35.4 (5.6) | 13 | **<0.001** |
| **Fasting glucose**, mmol/L | 5.4 (0.7) | 13 | 4.6 (0.4) | 13 | **0.044** |
| **Insulin**, IU/L | 18.3 (7.1) | 13 | 6.9 (3.6) | 11 | **<0.001** |
| **HOMA-IR** | 4.3 (1.5) | 13 | 1.4 (0.7) | 11 | **<0.001** |
| **Adiponectin**, μg/L | 5.2 (1.6) | 13 | 10.5 (4.1) | 11 | **0.003** |
| ApoM, mg/L | 6.8 (2.7) | 13 | 7.7 (2.7) | 13 | 0.190 |
| **Triglycerides**, g/L | 1.22 (0.52) | 13 | 0.70 (0.20) | 13 | **0.003** |
| **Total cholesterol**, g/L | 1.88 (0.28) | 13 | 1.81 (0.24) | 13 | **0.077** |
| **LDL-C**, g/L | 1.22 (0.21) | 13 | 1.09 (0.21) | 13 | **0.032** |
| **HDL-C**, g/L | 0.41 (0.09) | 13 | 0.56 (0.08) | 13 | **0.002** |
| **ApoA-I**, g/L | 1.43 (0.17) | 13 | 1.58 (0.19) | 13 | **0.002** |
| **ApoB-100**, g/L | 0.93 (0.17) | 13 | 0.84 (0.14) | 13 | **0.029** |
| ApoC-III, mg/L | 82.7 (41.9) | 13 | 74.1 (19.5) | 13 | 0.117 |
| **hs-CRP**, mg/L | 10.1 (8.0) | 13 | 2.3 (2.0) | 13 | **0.004** |
| **γ-GT**, IU/L | 34.6 (32.6) | 13 | 16.5 (14.2) | 12 | **0.017** |
| Creatinine, µmol/L | 63.7 (8.4) | 13 | 60.5 (2.9) | 12 | 0.099 |
| Systolic blood pressure, mm Hg | 115 (7) | 13 | 117 (14) | 13 | 0.202 |
| Diastolic blood pressure, mm Hg | 68 ± 14 | 13 | 73 ± 13 | 13 | 0.362 |
| eGFR, mL/min/1.73m^2^ | 109.6 (14.7) | 13 | 110.5 (14.3) | 12 | 0.461 |
| Data are expressed as means (SD).  Wilcoxon signed ranks test; *P*-values < 0.05 are indicated in bold.  ApoA-I: apolipoprotein A-I; ApoB-100: apolipoprotein B100; ApoC-III: apolipoprotein C-III; BMI, body mass index; eGFR, estimated glomerular filtration rate; hs-CRP, high-sensitivity C-reactive protein; γ-GT, gamma glutamyl transferase; HDL-C, high-density lipoprotein-cholesterol; HOMA-IR: Homeostasis Model Assessment of Insulin Resistance; LDL-C, low-density lipoprotein-cholesterol. | | | | | |

**Table S5. Anthropometric and clinical characteristics in the cohort C (n=267)**

|  | Mean (SD) |
| --- | --- |
| BMI, kg/m^2^ | 34.1 (4.7) |
| Fat mass, % body weight | 39.2 (7.6) |
| Waist circumference, cm | 106.3 (11.5) |
| Triglycerides, g/L | 1.37 (0.64) |
| Total cholesterol, g/L | 4.98 (0.99) |
| LDL-C, g/L | 3.10 (0.84) |
| HDL-C, g/L | 1.24 (0.34) |
| Fasting glucose, mmol/L | 5.2 (0.6) |
| Insulin, IU/L | 10.6 (4.8) |
| HOMA-IR | 2.7 (1.6) |
| hs-CRP, mg/L | 3.7 (3.2) |
| Systolic blood pressure, mm Hg | 126 (15) |
| Diastolic blood pressure, mmHg | 78 (11) |
| Data are expressed as means (SD).  BMI: body mass index; HDL-C: high-density lipoprotein cholesterol; hs-CRP: high-sensitivity C-reactive protein; HOMA-IR: Homeostasis Model Assessment of Insulin Resistance; LDL-C: low-density lipoprotein cholesterol. | |
